# Nationwide Trends and Outcomes in Major Gastrointestinal Cancer Surgery

**DOI:** 10.64898/2026.05.26.26354087

**Authors:** Rodolfo Eduardo de Andrade Espinoza, Leonardo S. L. Bastos, Silvio Hamacher, Jorge Ibrain Figueira Salluh, Fernando A. Bozza

**Affiliations:** Intensive Care Unit, Instituto Nacional do Câncer (INCA)/ Unidade II, Rio de Janeiro, Brazil; Department of Industrial Engineering (DEI), Pontifical Catholic University of Rio de Janeiro (PUC-Rio), Rio de Janeiro, RJ, 22451-900, Brazil; Tecgraf Institute, Pontifical Catholic University of Rio de Janeiro (PUC-Rio), Rio de Janeiro, RJ, 22451-900, Brazil; D’Or Institute for Research and Education (IDOR), Rio de Janeiro, RJ, 22281-100, Brazil; Oswaldo Cruz Foundation, Ministry of Health, Rio de Janeiro, Brazil; CHRC, NOVA Medical School, UNL, Lisboa, Portugal

**Keywords:** Gastrointestinal neoplasms, Surgical oncology, Surgical mortality, Health services research, Unified Health System

## Abstract

**Background:** Complex gastrointestinal (GI) oncologic surgeries carry substantial perioperative risk, and nationwide outcomes in low- and middle-income countries (LMICs) are underreported. This study aimed to evaluate national trends in surgical volume, in-hospital mortality, and intensive care unit (ICU) utilization for major GI cancer surgery in Brazil’s Unified Health System (SUS) over a 14-year period.

**Methods:** A population-based analysis was performed using national administrative databases to identify all adult patients undergoing colectomy, gastrectomy, pancreatic resection or esophagectomy for cancer in the SUS from 2010–2023. Annual rates were age-standardized according to the WHO standard population. Temporal trends were assessed using Poisson regression to estimate average annual percent change (AAPC) with 95% confidence intervals (CIs).

**Results:** A total of 179,337 hospital admissions were analyzed (median age 63 years; 48% female). Colectomies accounted for 72% of cases, followed by gastrectomies (19%), pancreatic resections (5%), and esophagectomies (3%). Although crude surgical volume increased, population-adjusted rates declined overall (AAPC –2.09%; 95% CI – 2.58 to –1.59), mainly due to reductions in gastrectomies and esophagectomies. Median hospital stay decreased from 9 to 7 days (AAPC –1.93%; 95% CI –2.79 to –1.06). Overall in-hospital mortality declined from 8.1% to 5.7% (AAPC –2.88%; 95% CI – 4.15 to –1.59). ICU utilization rose from 37% to 43% of admissions (AAPC +1.31%; 95% CI 0.91 to 1.71).

**Conclusion:** Over 14 years, in-hospital mortality and length of stay for major gastrointestinal cancer surgery declined within Brazil’s universal public health system. These temporal trends occurred alongside expansion of accredited oncology services and increased ICU utilization, although causal relationships cannot be established from administrative data. These findings should be interpreted as hypothesis-generating and highlight the need for more granular hospital-level data in LMIC settings.

## Introduction

Surgery is an essential intervention for the treatment of solid malignancies, particularly in early-stage. However, access to surgical care and the associated outcomes may vary substantially depending on the country’s socio-economic status (1–3). Global studies have consistently shown that postoperative increased mortality rates in low- and middle-income countries (LMICs), mainly in cancer surgery (4). This disparity has been attributed to several factors, including the late diagnosis (locally advanced or metastatic disease at presentation), hospital resource constraints, shortages of adequately trained perioperative care teams, and failures in the ability to manage surgical complications (5).

In LMICs, data on the burden of cancer surgery, postoperative complications and hospital outcomes remain scarce. When available, such data often come from single institutions or small regional cohorts, limiting their generalizability. Consequently, the generation of granular yet nationally representative data is critical to enable the development of equitable public health policies and sustainable improvements in surgical care systems.(6,7).

In Brazil, an upper middle-income country with universal healthcare coverage through the Unified Health System (Sistema Único de Saúde – SUS), the National Policy for Cancer Care was established in 2005, defining the criteria for the accreditation of specialized oncology services (8). However, knowledge regarding the volume of complex oncologic surgeries and their hospital outcomes remains limited.

The present study aims to describe the volume and outcomes of complex oncologic surgeries performed in the SUS between 2010 and 2023 and analyze temporal trends in in-hospital mortality.

## Materials and Methods

### Data Collection

Data was extracted from two databases that are available for public access. The first dataset, SIHSUS (Hospitalar Admission System) (9) was obtained directly from the DATASUS website (https://datasus.saude.gov.br/). DATASUS is the Department of Information and Informatics of the Brazilian Unified Health System, Ministry of Health, Brazil. The SIHSUS gathers the Authorizations for Hospitalization (AIH–Autorização de Internação Hospitalar). Each AIH registry contains de-identified data on inpatient demographic characteristics (age and gender), use of intensive care unit, diagnosis at admission (ICD-10), hospital main procedure, length of stay, patient outcome (discharge from hospital or death) and other information. The populational dataset was obtained from the Brazilian Institute of Geography and Statistics (IBGE—Instituto Brasileiro de Geografia e Estatística) for each year, age, and gender, after the Brazilian Census (10). We analyzed anonymized public datasets containing patients. All data were aggregated before access and analysis. Consequently, the ethics principles were respected, and informed consent was waived.

#### Study population

In the last census (2022), Brazil had a population of 203,080,756, compared to 193,543,969 in 2009. Of these, more than 75% depends exclusively on the SUS for health services.

We analyzed colectomies, gastrectomies, pancreatic resections and esophagectomies. All hospital admissions for each of the four groups of procedures analyzed in our study were identified using the appropriate codes from the SUS Procedure, Medication, Orthotics, Prosthetics, and Special Materials Management System (SIGTAP) (11). The codes are provided in Supplement 1. These procedures were selected because they are widely recognized as high-risk oncologic operations associated with substantial perioperative mortality and resource utilization (12,13). In studies conducted predominantly in high-income countries, they have also served as important indicators of perioperative care capacity, institutional infrastructure, and overall surgical system performance (3,13,14). Additionally, we selected the procedures according to International Classification of Diseases, 10th Revision (ICD-10) neoplasm codes : C15 (esophageal cancer), C16 (gastric cancer), C24 (biliary tract cancer), C25 (pancreas cancer), C18, C19, C20 and C21 (colon and rectum cancer), C26 (undefined digestive organ cancer) and D13 (benign digestive organ neoplasm). Only AIHs from adult patients were analyzed (older than 20 years old).

The primary aim was to evaluate the overall number of hospital admissions and in-hospital deaths. Secondary endpoints were the hospitalization rate per 100,000 population, age, and ICU admissions. We obtained rates per 100,000 population using direct standardization data from the WHO standard population distribution for 2000-2025 (15), and the estimates of the adult Brazilian population using IBGE as the reference for the study population.

The Annual Average Percent Change (AAPC) and its associated 95% confidence interval (95% CI) were computed to assess the evolution of outcomes throughout the study period. AAPC offers a reliable assessment of the annual average trend of increase (positive AAPC) or reduction (negative AAPC). The AAPC was calculated using log-linear Poisson regression, considering the year as the single predictor (the reference year was 2010) for the crude number of hospital admissions and in-hospital deaths. We added the age-adjusted denominator to the Poisson model for age-standardized rates as an offset variable. We calculated the AAPC from 2010 to 2023, as well as the percentage change (%Δ) over these periods. All analyses were performed using Python 3.10.4, with packages Plotly, *Pandas, NumPy, Statsmodel, Seaborn*, and *Matplotlib*.

## Results

From January 1st 2010 to December 31st 2023, 164,390,800 hospital admissions were identified and after the selection of surgical procedures, ICD-10 and age, 179,337 admissions were analyzed (Fig 1).

**Figure 1.**
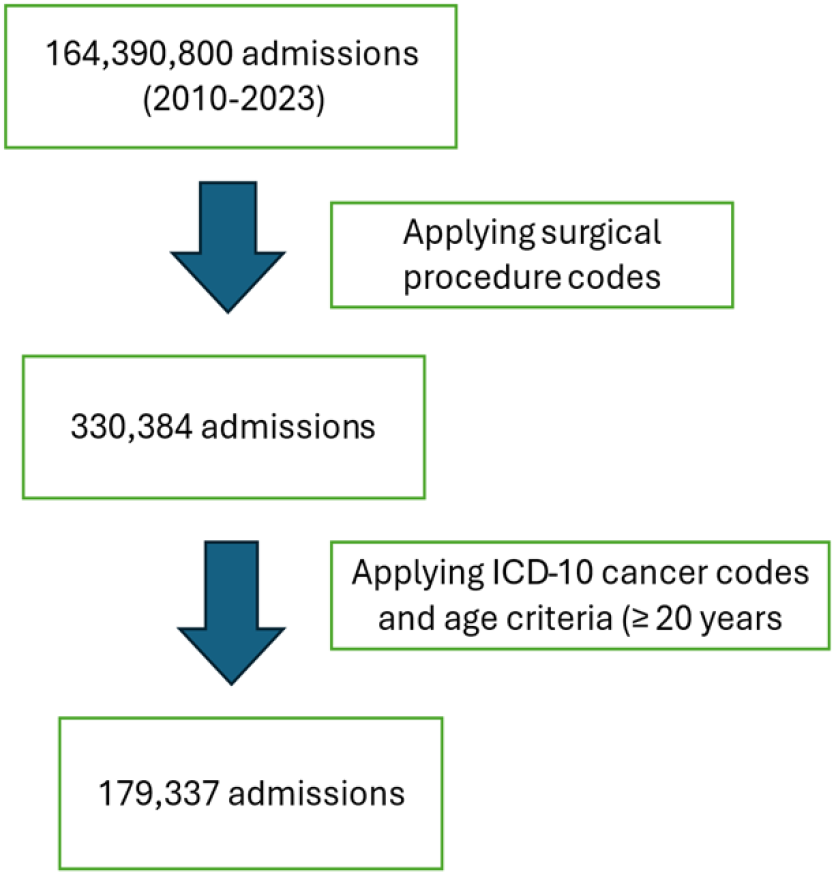
Flowchart of the study population. ICD-10: International Classification of Diseases, 10th Revision Age ≥ 20 years

The median age was 63 years (IQR 54-71), the female gender 48%, the hospital length of stay 8 days (IQR 5-12), and the hospital mortality rate 7.1%. ICU admission was required by 40% of the patients, with an ICU LOS 2 days (IQR2-4) and 13% mortality (Table 1).

**Table 1.** Baseline characteristics of all surgical admissions, 2010–2023.

| Characteristic | 2010, N<br>= 12,499 | 2011, N<br>= 12,045 | 2012, N<br>= 11,726 | 2013, N<br>= 12,548 | 2014, N<br>= 12,373 | 2015, N<br>= 12,253 | 2016, N<br>= 12,179 | 2017, N<br>= 12,617 | 2018, N<br>= 13,357 | 2019, N<br>= 13,894 | 2020, N<br>= 12,578 | 2021, N<br>= 13,060 | 2022, N<br>= 13,576 | 2023, N<br>= 14,632 | Overall, N<br>= 179,337 |
| --- | --- | --- | --- | --- | --- | --- | --- | --- | --- | --- | --- | --- | --- | --- | --- |
| Age (years) | 62 (52, 71) | 62 (52, 71) | 62 (53, 71) | 63 (53, 71) | 63 (53, 71) | 63 (54, 71) | 63 (54, 71) | 63 (54, 71) | 63 (54, 71) | 63 (55, 71) | 63 (54, 71) | 64 (55, 72) | 64 (55, 71) | 64 (55, 72) | 63 (54, 71) |
| Female, n (%) | 5,744 (46%) | 5,447 (45%) | 5,484 (47%) | 5,911 (47%) | 5,906 (48%) | 5,895 (48%) | 5,891 (48%) | 6,102 (48%) | 6,548 (49%) | 6,819 (49%) | 6,070 (48%) | 6,446 (49%) | 6,851 (50%) | 7,354 (50%) | 86,468 (48%) |
| Surgery, n (%) |  |  |  |  |  |  |  |  |  |  |  |  |  |  |  |
| Colectomies | 8,186 (65%) | 7,952 (66%) | 7,915 (67%) | 8,844 (70%) | 8,850 (72%) | 8,846 (72%) | 8,752 (72%) | 9,185 (73%) | 9,715 (73%) | 10,199 (73%) | 9,420 (75%) | 9,823 (75%) | 10,423 (77%) | 11,344 (78%) | 129,454 (72%) |
| Gastrectomies | 3,116 (25%) | 2,952 (25%) | 2,755 (23%) | 2,518 (20%) | 2,404 (19%) | 2,348 (19%) | 2,333 (19%) | 2,393 (19%) | 2,568 (19%) | 2,563 (18%) | 2,214 (18%) | 2,258 (17%) | 2,206 (16%) | 2,247 (15%) | 34,875 (19%) |
| Pancreatic Resections | 497 (4.0%) | 518 (4.3%) | 545 (4.6%) | 654 (5.2%) | 658 (5.3%) | 621 (5.1%) | 673 (5.5%) | 680 (5.4%) | 713 (5.3%) | 758 (5.5%) | 674 (5.4%) | 684 (5.2%) | 687 (5.1%) | 747 (5.1%) | 9,109 (5.1%) |
| Esophagectomies | 700 (5.6%) | 623 (5.2%) | 511 (4.4%) | 532 (4.2%) | 461 (3.7%) | 438 (3.6%) | 421 (3.5%) | 359 (2.8%) | 361 (2.7%) | 374 (2.7%) | 270 (2.1%) | 295 (2.3%) | 260 (1.9%) | 294 (2.0%) | 5,899 (3.3%) |
| ICU Admissions n, (%) | 4,570 (37%) | 4,379 (36%) | 4,361 (37%) | 4,819 (38%) | 4,850 (39%) | 4,906 (40%) | 4,865 (40%) | 5,002 (40%) | 5,437 (41%) | 5,912 (43%) | 5,075 (40%) | 5,044 (39%) | 5,561 (41%) | 6,267 (43%) | 71,048 (40%) |
| In-hospital death, n (%) | 1,045 (8.4%) | 988 (8.2%) | 886 (7.6%) | 1,051 (8.4%) | 980 (7.9%) | 947 (7.7%) | 881 (7.2%) | 826 (6.5%) | 874 (6.5%) | 882 (6.3%) | 834 (6.6%) | 884 (6.8%) | 770 (5.7%) | 838 (5.7%) | 12,686 (7.1%) |
| Hospital LOS, median<sup>1</sup> (IQR) | 9 (7, 14) | 9 (6, 14) | 9 (6, 14) | 9 (6, 14) | 9 (6, 14) | 9 (6, 14) | 8 (6, 13) | 8 (6, 12) | 8 (5, 12) | 8 (5, 12) | 7 (5, 11) | 7 (5, 10) | 7 (5, 10) | 7 (5, 10) | 8 (5, 12) |
| In-hospital death from ICU, n (%) | 705 (15%) | 673 (15%) | 591 (14%) | 722 (15%) | 690 (14%) | 684 (14%) | 639 (13%) | 553 (11%) | 641 (12%) | 609 (10%) | 619 (12%) | 636 (13%) | 595 (11%) | 644 (10%) | 9,001 (13%) |
| ICU LOS, median<sup>1</sup>(IQR) | 2 (1, 5) | 2 (1, 4) | 2 (1, 4) | 3 (2, 5) | 3 (1, 5) | 3 (2, 4) | 3 (2, 5) | 3 (2, 4) | 3 (2, 4) | 2 (2, 4) | 2 (1, 4) | 2 (2, 4) | 2 (2, 4) | 3 (2, 4) | 2 (2, 4) |

<sup>1</sup>Median (IQR=interquartile range); n (%); LOS : Length of stay in days

A total of 129,454 colorectal surgeries, 34,875 gastrectomies, 9,109 pancreatic resections, and 5,899 esophagectomies were performed during the study period.Although the total crude number of admissions increased, the AAPC (adjusted per 100,000) was negative (AAPC: −2.09% (−2.58, −1.595)). The result was more pronounced in esophagectomies −9.23% (−10.61 to −7.821), gastrectomies −4.99% (−5.83 to −4.13) and colorectal surgeries −1.26% (−1.684 to −0.838). Conversely, pancreatic resections showed a slight positive AAPC: +0.17% (−1.10, +1.45) (Fig2).

**Figure 2.**
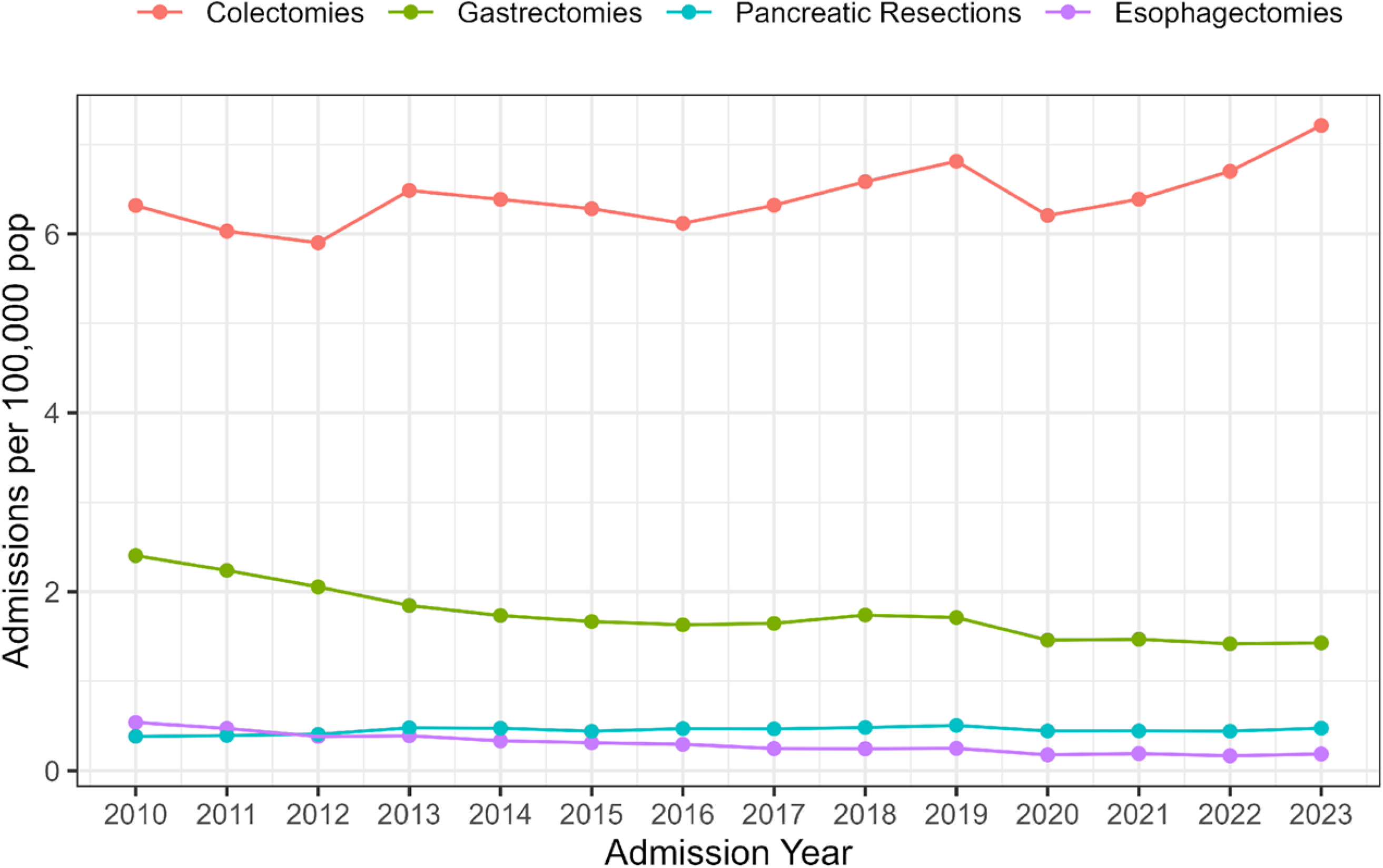
Annual admissions per 100,000 population for colectomies, gastrectomies, pancreatic resections, and esophagectomies performed in the Brazilian Unified Health System (SUS).

Overall, the hospital length of stay decreased over time, from 9 days (IQR 7– 14) in 2010 to 7 days (IQR 5–10) in 2023, with an AAPC of –1.93% (95% CI: –2.79 to–1.06). When stratified by type of surgical procedure, a similar trend was observed: colectomies –1.79% (–2.28 to –1.29), gastrectomies –1.94% (–2.68 to –1.21),pancreatic resections –2.21% (–3.00 to –1.42), and esophagectomies –1.76% (–2.44 to –1.09) (Table 1).

The general in-hospital mortality rates decreased from 8.1% in 2010 to 5.7% in 2023 (table 1 and figure 3), with an AAPC of –2.88% (95% CI: –4.15 to –1.59). All procedures demonstrated a downward trend in mortality: colectomies AAPC –2.43% (95% CI: –4.22 to –0.60), gastrectomies –2.56% (95% CI: –5.56 to 0.54), pancreatic resections AAPC –3.26% (–7.40 to –1.06), and esophagectomies –1.64% (–7.22 to 4.25) (table 1 and figure 4).

**Figure 3.**
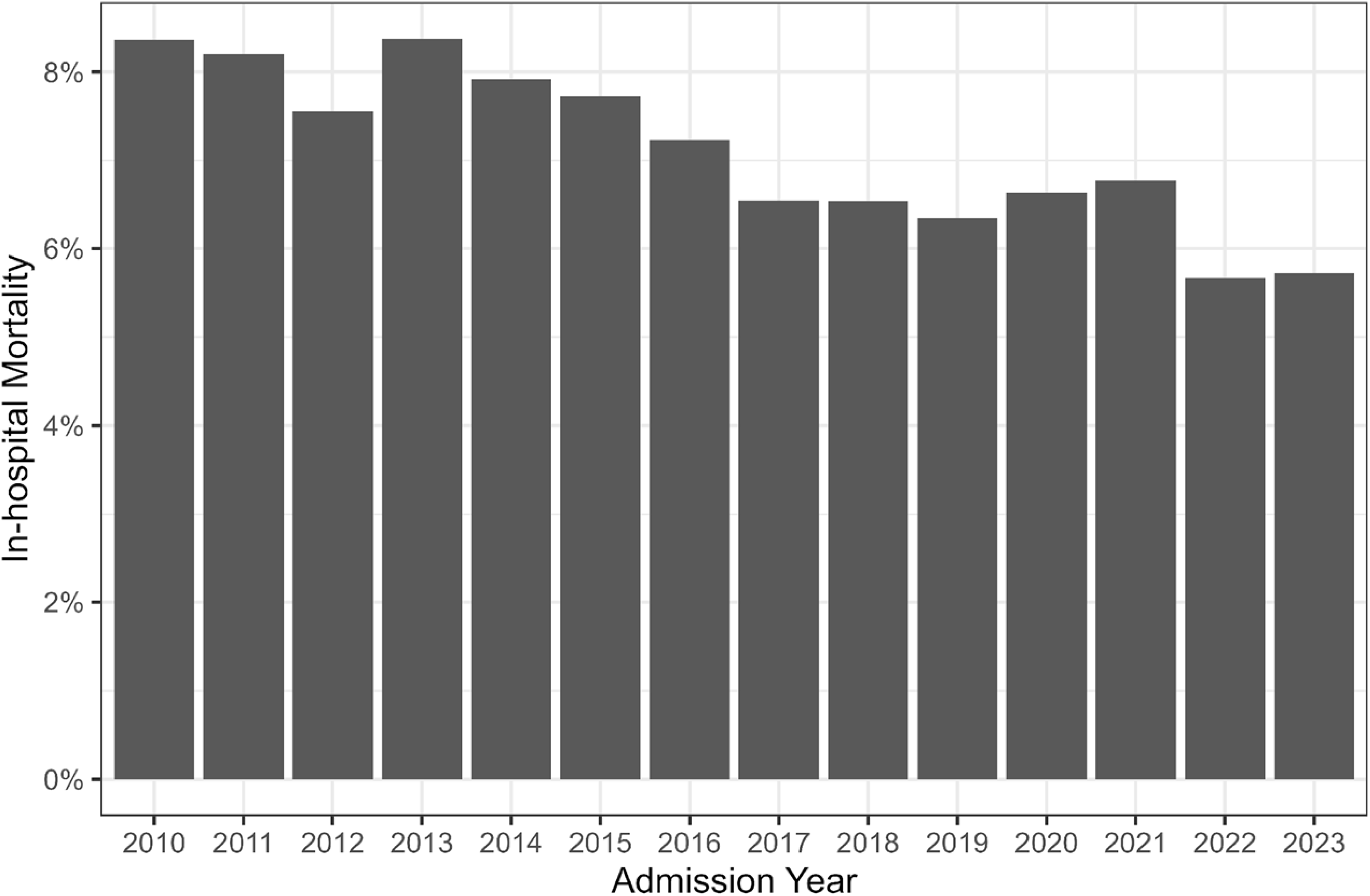
Overall in-hospital mortality(%) for all included surgical procedures performed in the Brazilian Unified Health System (SUS), 2010-2023

**Figure 4.**
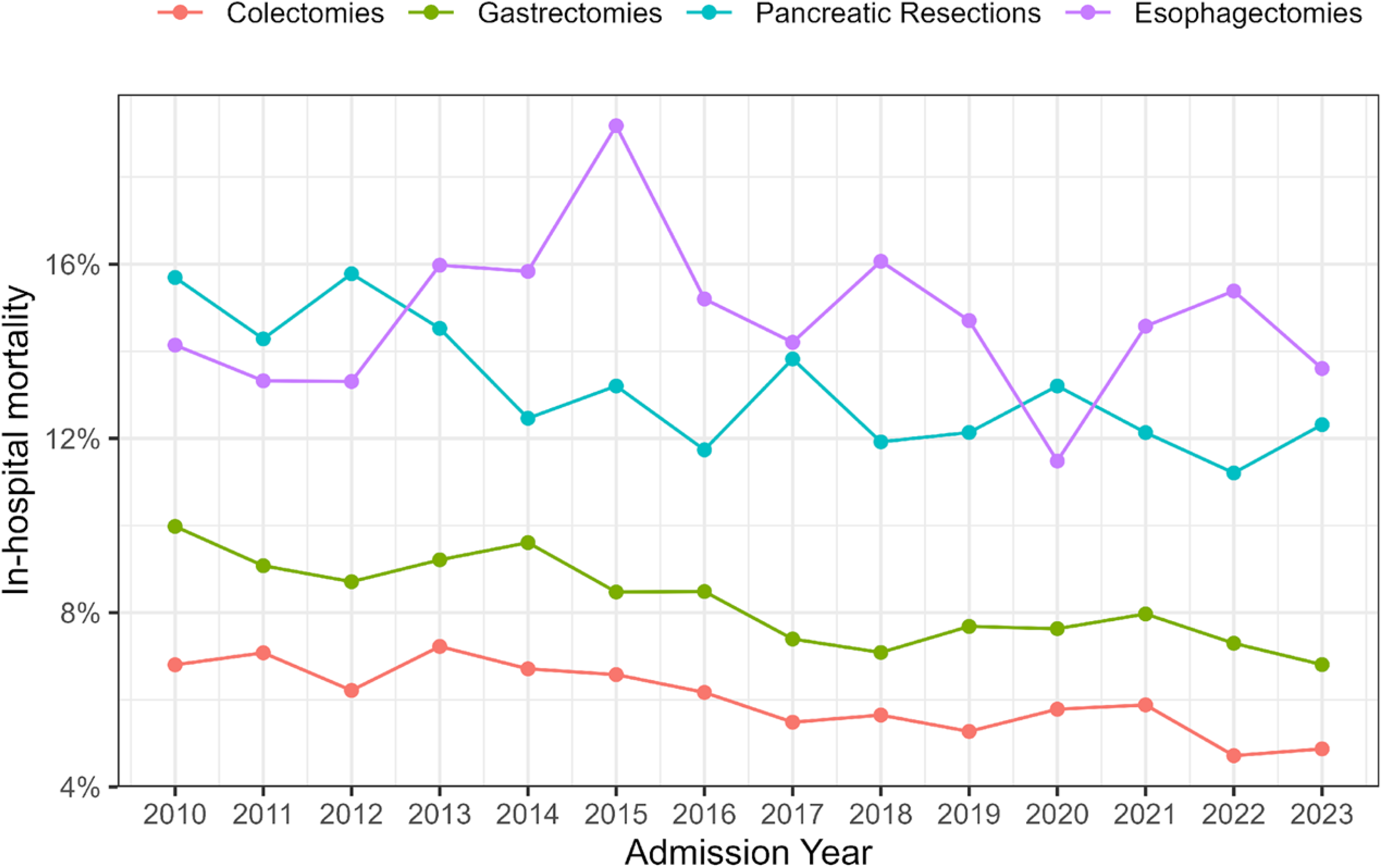
Annual in-hospital mortality (%) for colectomies, gastrectomies, pancreatic resections and esophagectomies performed in the Brazilian Unified Health System (SUS), 2010-2023.

The ICU admissions increased during the study period. Observing all surgeries, from 4,570 (37%) admissions in 2010 to 6,267 (43%) in 2023 with AAPC + 1.31% (0.91, 1.71). When analyzing ICU admissions by type of surgery, an increasing trend was observed for all procedures except for esophagectomies : colectomies +0.874% (1.38 to 2.37), gastrectomies +1.554% (0.76 to 2.36), pancreatic resections +0.739% (95% CI: 0.10 to 1.37), and esophagectomies +1.679% (–0.02 to 3.41) (table 1 and figure 5).

**Figure 5.**
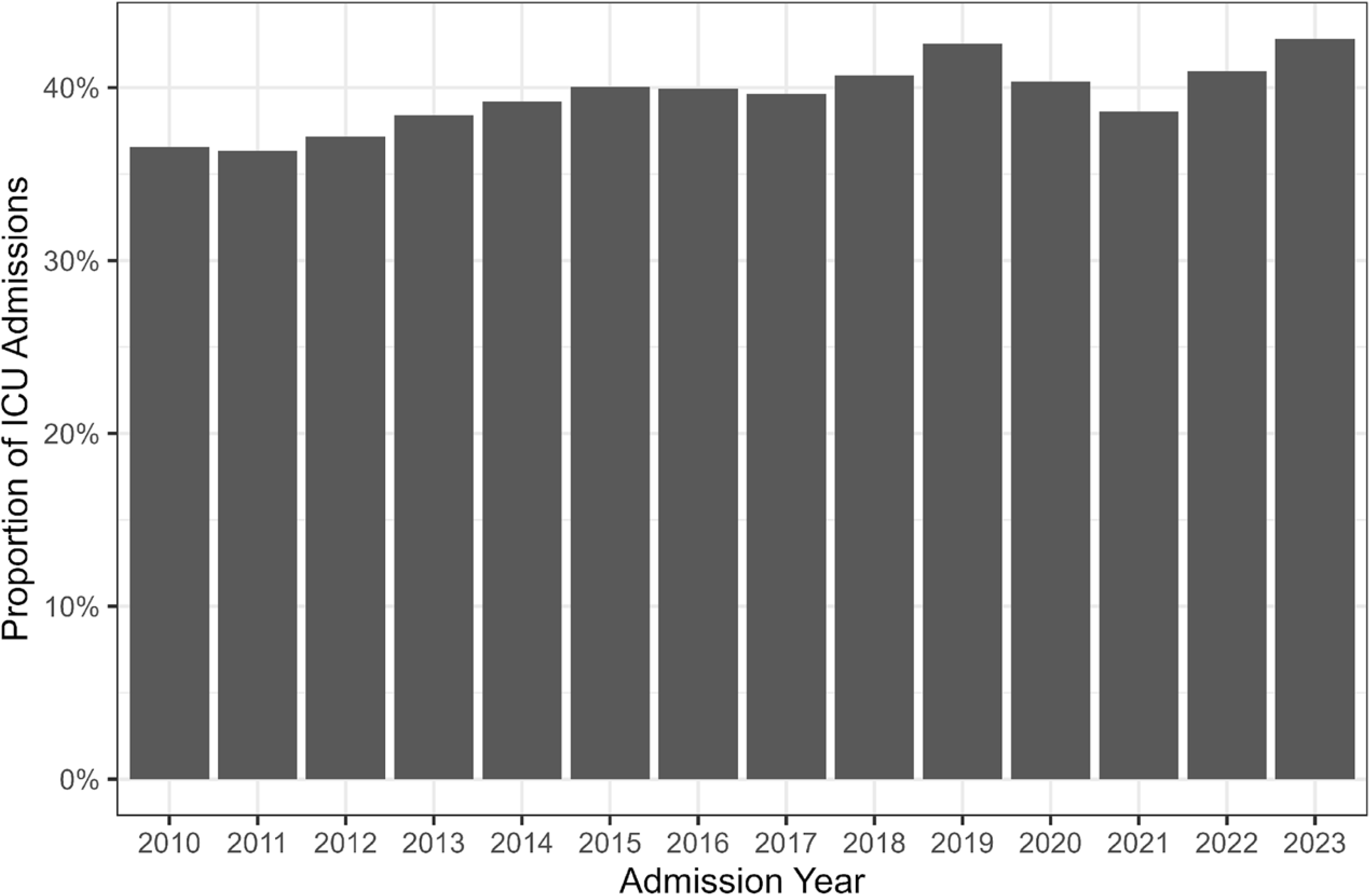
Proportion of patients admitted to intensive care following the analyzed surgeries in the Brazilian Unified Health System (SUS).

## Discussion

This nationwide, population-based analysis of complex gastrointestinal oncologic surgeries in the Brazilian public health system over a 14-year period, revealed a consistent decline in hospital mortality, accompanied by reductions in length of stay and increases in intensive care utilization. While crude surgical volumes increased slightly, population-adjusted rates decreased, particularly for gastrectomies and esophagectomies, whereas pancreatic resections remained stable. These findings offer valuable insight into surgical oncology performance in an upper middle-income country.

The variation in adjusted surgical rates cannot be attributed solely to changing cancer incidence patterns. Although gastric and esophageal cancers are decreasing, colorectal and pancreatic cancer cases have risen in Latin America (16–18). However, in our context, demographic growth and persistent inequities in service distribution likely compound these trends (6). Previous research has documented geographic disparities in cancer bed distribution and surgical capacity within Brazil (8), implying that service availability remains a limiting factor despite nominal increases in accredited centers.

Mortality rates observed in our study are comparable to those reported in other LMICs(2,19). Nonetheless, the observed mortality rates remain above the most recent global benchmarks for perioperative outcomes in major cancer surgery, with the discrepancy being most pronounced for esophagectomies (4,20,21). The higher mortality associated with major oncologic surgery in LMICs can be attributed to several factors. Surgical treatment of more advanced neoplasms is linked to increased perioperative mortality(22), a scenario frequently encountered in health systems with limited access to timely care often observed in cancer care in LMIC. Differences in perioperative support, availability of critical care resources, institutional infrastructure, coordinated care pathways, and multidisciplinary surgical expertise may further contribute to the marked variation in postoperative mortality observed across healthcare systems, particularly in resource-constrained settings (23).

The mechanisms underlying the observed reduction in mortality cannot be directly evaluated using administrative data. Nevertheless, the temporal trends observed in this study occurred alongside expansion of accredited oncology services and increased ICU utilization within the Brazilian public health system. Brazil expanded the number of accredited cancer centers from 349 in 2010 to 547 in 2023, reflecting broader efforts to strengthen oncologic care delivery nationwide. Previous studies from high-income settings have suggested that specialized perioperative infrastructure and multidisciplinary care may be associated with improved surgical outcomes (24–26). However, the present study was not designed to determine whether these factors directly contributed to the observed mortality trends.

The present study has some strengths. nationwide scope, capturing all publicly funded high-complexity gastrointestinal cancer surgeries over more than a decade, using standardized ICD-10 and procedure coding. The large sample size and age-standardized rates allow for robust temporal trend analysis. However, there are also limitations including reliance on administrative data, absence of clinical details such as cancer stage, comorbidities, and specific postoperative complications, and exclusion of the private sector, which accounts for a smaller proportion of cancer surgeries in Brazil. Potential coding errors and unmeasured confounders are inherent to secondary database analyses. Moreover, the database lacks key clinical and process-related variables that could help elucidate some observed trends, such as the decline in mortality and length of stay, or the increased use of intensive care units.

Linking administrative datasets with cancer registries and mortality systems could enable richer risk adjustment and outcome profiling, bridging gaps in evidence for LMIC surgical oncology. Prospective multicenter collaborations would further contextualize perioperative mortality and complication rates, supporting targeted quality improvement initiatives. Further research is needed to determine whether the expansion of specialized cancer centers has contributed to the observed reduction in mortality.

## Conclusion

Over a 14-year period, major gastrointestinal cancer surgery within the Brazilian public health system was associated with progressive reductions in in-hospital mortality and length of stay, despite declining population-adjusted surgical rates. These findings occurred alongside expansion of accredited oncology services and greater use of intensive care resources during the study period, although the underlying mechanisms cannot be determined from administrative data alone. This nationwide analysis provides important insight into surgical oncology outcomes in an upper middle-income country and highlights the need for more granular hospital-level and clinical data in LMIC settings.

## Data Availability

The data underlying the results presented in this study are publicly available from the Brazilian Unified Health System (SUS) Hospital Information System (SIH-SUS) through the DATASUS platform of the Brazilian Ministry of Health. Data can be accessed at: datasus.saude.gov.br

https://datasus.saude.gov.br/acesso-a-informacao/producao-hospitalar-sih-sus/

